# Differential risk of healthcare workers versus the general population during outbreak, war and pandemic crises

**DOI:** 10.1101/2024.05.30.24308231

**Authors:** John P.A. Ioannidis

## Abstract

Healthcare workers may have different risk for severe outcomes compared with the general population during diverse crises. This paper introduces the concept of healthcare worker versus population hazard (HPH), the risk of an outcome of interest in active healthcare workers compared with the general population they serve. HPH can be expressed with relative risk (HPH(r)) and absolute risk difference (HPH(a)) metrics. Illustrative examples are drawn from infectious outbreaks, war, and the COVID-19 pandemic on death outcomes. HPH can be extreme for lethal outbreaks (HPH(r)=30 to 143, HPH(a)=8 to 91 per 1000 for Ebola deaths in 3 Western African countries in 2013-5), and modestly high in relative terms and very high in absolute terms for protracted, major armed conflicts (HPH(r)=1.38 and HPH(a)=10.2 for Syria during 2011-2024). Conversely, healthcare workers had 8-12-fold lower risk than the population they served for pandemic excess deaths (physicians in USA) or COVID-19 deaths (physicians in Ontario, healthcare workers in Finland), while healthcare workers in Indonesia did not have this advantage for COVID-19 deaths versus the general population. HPH is susceptible to data inaccuracies in numbers of at-risk populations and of outcomes of interest. Importantly, inferences about healthcare worker risk can be misleading, if deaths of retired healthcare workers contaminate the risk calculations – as in the case of misleading early perceptions of exaggerated COVID-19 risk for healthcare professionals. HPH can offer useful insights for risk assessment to healthcare professionals, the general public, and policy makers and may be useful to monitor for planning and interventions during crises.

## INTRODUCTION

Healthcare workers face numerous infectious and other hazards in their practice [1–3]. Specific hazards may arise or get particularly exacerbated, noted and feared during crises, such as infectious outbreaks, wars, and pandemics. Hazards may affect both the healthcare workforce and the general population. It would be useful to have measures that quantify the risk of a severe outcome, e.g. deaths related to the crisis, in active healthcare professionals versus the population that they serve. Healthcare professionals may be at higher risk than the general population, if they are more exposed (e.g. to an infectious agent at work) or even targeted (e.g. in armed conflicts where militants target specifically healthcare professionals and facilities) [4]. Conversely, healthcare professionals may be at relatively lower risk, if they have favorable demographic or other features (e.g. healthy worker effect) or better access to effective preventive and therapeutic interventions.

This article introduces the concept of the healthcare worker versus population hazard (HPH) with metrics of relative risk (HPH(r)) and absolute risk difference (HPH(a)) that are applicable across diverse types of crises. HPH metrics can demonstrate how much higher (or lower) the risk of a serious outcome is for active healthcare professionals versus the general population they serve. HPH is exemplified with data from outbreaks, war, and the COVID-19 pandemic. HPH(r) and HPH(a) information may allow healthcare workers to calibrate the risk that they face given their professional activity; and may facilitate the general public to better understand whether their health providers are at much higher (or lower) risk than they are. Policy makers may also use this information to make decisions that minimize risks to healthcare workers and preserve this valuable workforce that is in short supply in many settings around the world [5] – with shortages often becoming further pronounced during crises.

## METHODS

### Definitions and conceptual issues

HPH is the differential risk of a serious outcome of interest among active healthcare professionals versus among the general population they serve. It can be presented with metrics of relative risk (HPH(r)) and absolute risk difference (HPH(a)). All examples used here focus on mortality outcomes (death due to specific causes or excess death estimates), but HPH can be applied to any outcome. Furthermore, all examples used here pertain to the healthcare workforce and general population at the level of whole countries or large provinces. All healthcare professionals in a given country or large province cumulatively serve the entire population therein. However, substantial inequalities may exist within any country/province, both in the availability of healthcare workers and in access to them by different population groups. Therefore HPH metrics should be seen as average estimates. The same principle can be applied also to localized geographic areas and to specific communities and groups; or, conversely, can be expanded to many countries or even the whole world. As crises evolve over time, relative and absolute risks may also increase or decrease disproportionately in healthcare workers versus in the communities they serve.

HPH metrics should not be confused with standardized metrics like proportional mortality ratios or standardized mortality ratios. In these ratios, one group of interest (e.g. healthcare workers) is compared against a reference population after adjustments for demographics, or even for additional variables, such as socioeconomic status, to achieve comparability. HPH metrics make no such adjustments, since these adjustments might explain away some of the differential risk between healthcare professionals and the general population.

HPH estimates will be biased, if the determination of the outcome of interest (e.g. death from some specific cause) is differentially affected in the healthcare workers group versus in the general population. Sometimes healthcare workers have more complete documentation of some causes of death versus the general population. E.g., COVID-19 deaths are undercounted in countries with suboptimal death registration [6] and correction for undercounting would be appropriate. Undercounting may not exist, or may be much smaller in active healthcare professionals, due to the nature of their work and/or use of comprehensive, specialized registries. An illustrative example of this correction is applied for COVID-19 deaths in Indonesia (see below).

HPH estimates will also need to be corrected, if during a crisis, healthcare workers flee the country at more massive rates that the general population. E.g. in armed conflicts [7], health professionals may have better means to immigrate and/or an extra urge to flee if they witness that they are targeted by militants. An illustrative example attempting to correct for reduced healthcare worker numbers was applied in the Syria armed conflict (see below).

HPH metrics may also be calculated for healthcare professionals of different age groups or as defined by other characteristics (e.g. specific healthcare profession or specialty, type and intensity of exposure, etc.). An illustrative example is provided here on pandemic excess deaths in the USA (see below) [8].

### Data and analyses

The following illustrative examples are dissected here with estimation of HPH metrics: Western African Ebola outbreak in Guinea, Liberia and Sierra Leone; armed conflict in Syria; and COVID-19 pandemic (USA, Ontario, Finland, Indonesia). The examples are selected to cover diverse types of crises with very different HPH estimates. Effects are calculated as HPH(r)=(health care worker deaths/healthcare worker population)/(general population deaths/general population) and HPH(a)=(health care worker deaths/healthcare worker population)-(general population deaths/general population).

For Ebola, information on overall and healthcare workers’ deaths due to the infection during 2013-2015 in Guinea, Liberia and Sierra Leone was derived from a WHO situational report [9]. Total numbers of healthcare workers were derived from CDC reports for Guinea [10] and Sierra Leone [11] and a technical report for Liberia [12]. Populations for these countries in 2013 was obtained from [13].

For the armed conflict in Syria, information on total deaths due to the armed conflict since 2011 was derived from the Syrian Observatory for Human Rights [14] and on deaths of physicians due to the armed conflict was derived from Physicians for Human Rights [15]. Reliable information on the total number of healthcare workers is not available, while the number of physicians is available for pre-crisis levels (2009) [7]. However, most physicians fled the country during the armed conflict [15]. Therefore, the calculations assumed a correction factor of 4, assuming that the effective number of physicians in Syria averaged only a quarter the pre-crisis levels. Data on deaths of all medical personnel are available also on an annual basis allowing HPH calculation per calendar year in a supplementary analysis (Supplementary text). The Syria population in each year was obtained from UN Population Division [16] and is also an approximation given the substantial uncertainty in number of refugees.

For USA excess deaths of active physicians during the two pandemic years 2020-2021, data were derived from a previous publication that used the archives of the American Medical Association Masterfile and the corresponding Deceased Physician File [8]. That publication provided excess death estimates only for physicians 45-84 years old, since younger strata had very low death numbers, but also offered data for granular age strata (45-64, 65-74, 75-84 years) and according to whether physicians provided direct patient care or not. An excess death estimate for the USA population in 2020-21 was obtained as proposed by Levitt et al. using age-adjustment [17].

Data on COVID-19 deaths in active Ontario physicians until end-2022 and the number of active Ontario physicians were derived from a previous analysis [18] along with communication with the authors to clarify deaths occurring within 30 days after COVID-19. Population data were obtained from [19] and Ontario COVID-19 deaths until end-2022 were obtained from [20].

Data on COVID-19 deaths in healthcare workers in Finland until mid-2021 and the number of registered healthcare workers in Finland were derived from a previous national registry analysis [21]. Population data were obtained from [22] and Finland COVID-19 deaths until mid-2021 were obtained from [23].

For Indonesia, data on COVID-19 deaths in healthcare workers (based on an available specialized registry) and on the entire population (based on available data from the ministry of health) and data on the country population and the number of healthcare workers (from the ministry of health) were already compiled in [24]. Given that COVID-19 deaths in the general population were substantially under-reported in Indonesia, HPH calculations used a corrected estimate of total COVID-19 deaths in the same period [25]. Sensitivity analyses examined the impact of 2- and 3-fold undercounting of COVID-19 deaths also among healthcare workers.

## RESULTS

### Datasets

Table 1 shows the relevant data for the illustrative examples. All examples pertain to cause-specific deaths (Ebola, war, COVID-19), except for USA in 2020-2021 where excess deaths (a composite of multiple causes of death affected by COVID-19 and the measures taken) are considered. All healthcare workers were considered in the Ebola outbreaks and in COVID-19 deaths in Finland and Indonesia. In other examples (Syria war, COVID-19 in USA and Ontario), active physicians were analyzed.

**Table 1.**
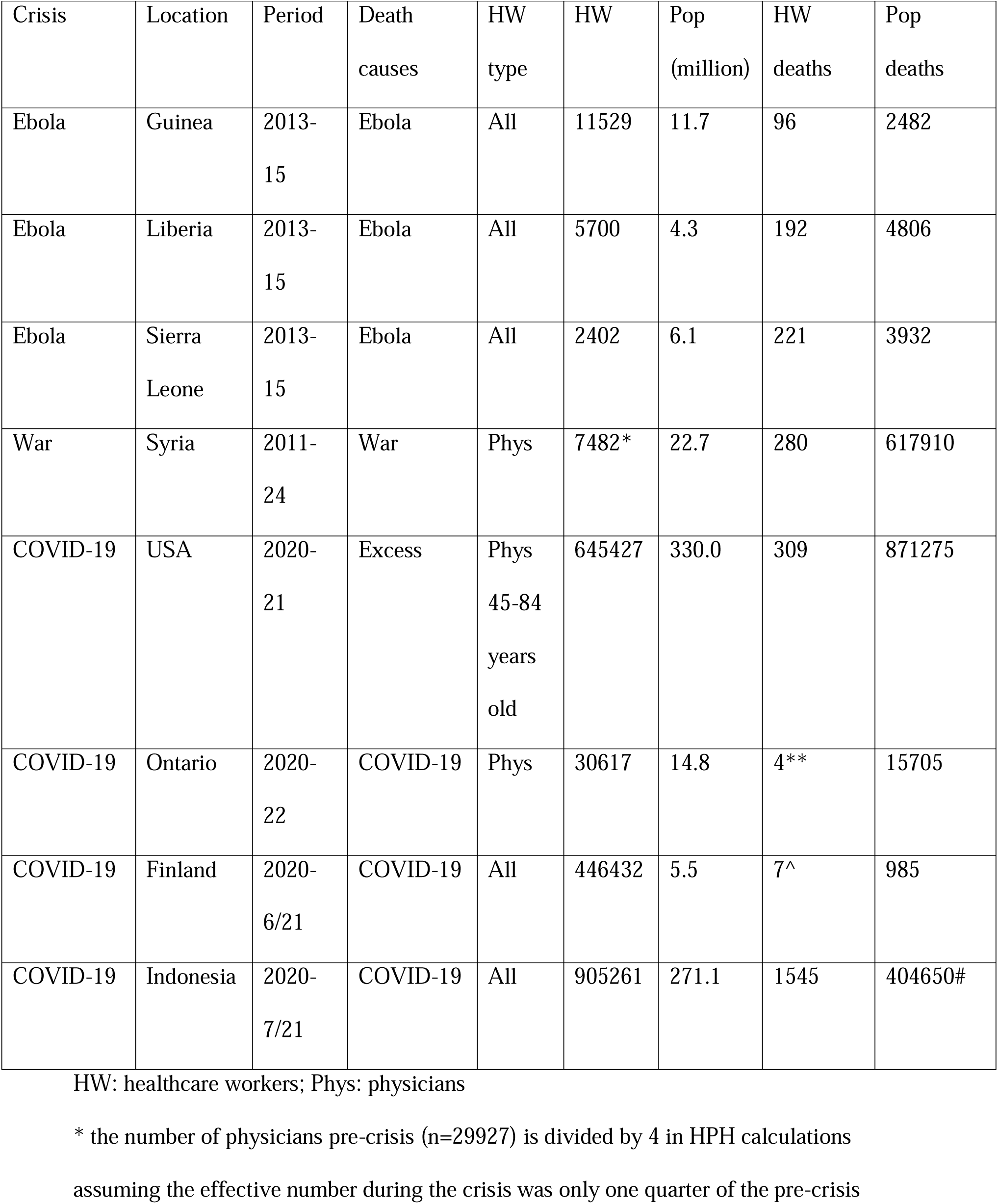
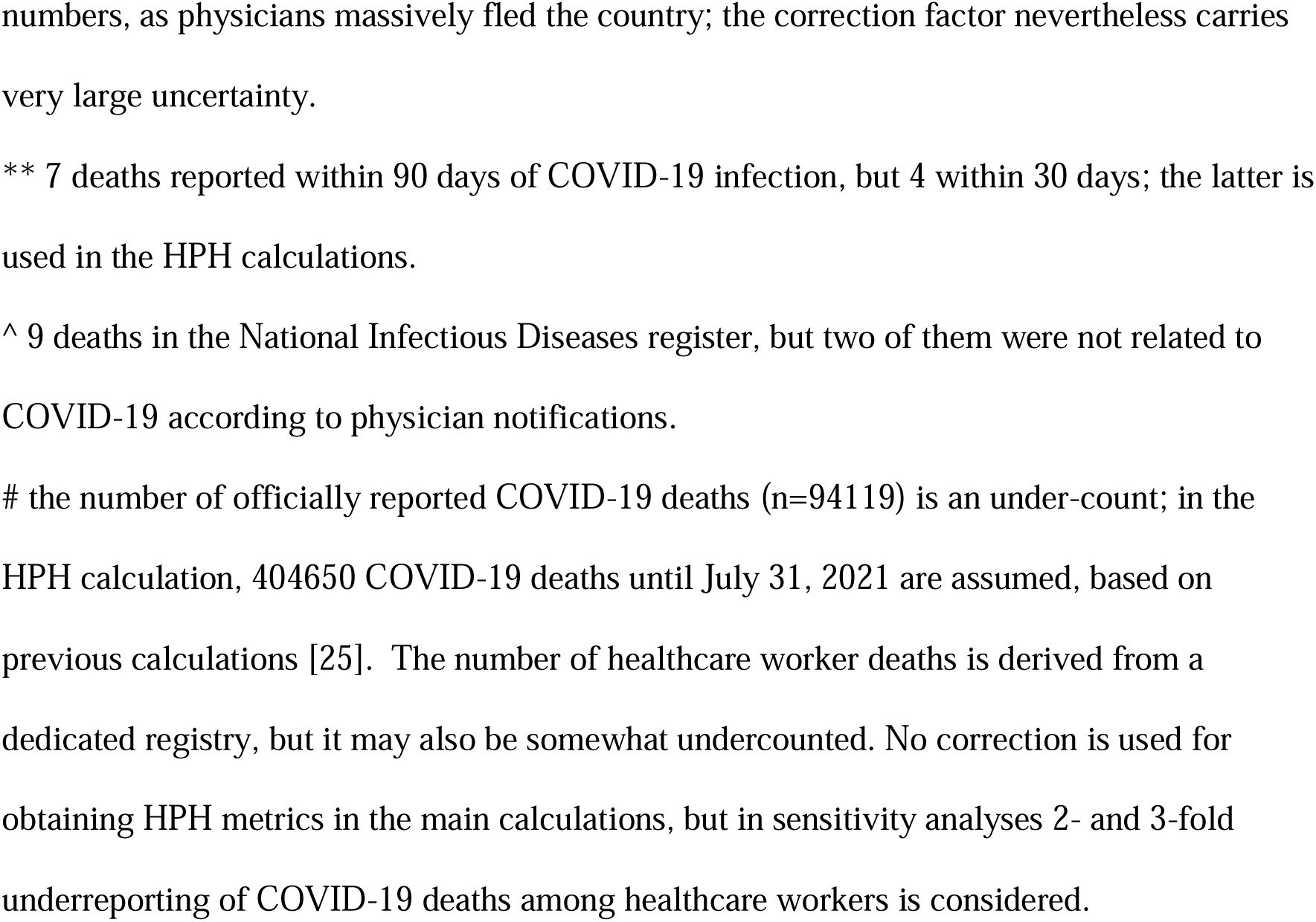
Data on healthcare workers and populations and mortality outcomes in illustrative examples.

| Crisis | Location | Period | Death causes | HW type | HW | Pop (million) | HW deaths | Pop deaths |
| --- | --- | --- | --- | --- | --- | --- | --- | --- |
| Ebola | Guinea | 2013-15 | Ebola | All | 11529 | 11.7 | 96 | 2482 |
| Ebola | Liberia | 2013-15 | Ebola | All | 5700 | 4.3 | 192 | 4806 |
| Ebola | Sierra Leone | 2013-15 | Ebola | All | 2402 | 6.1 | 221 | 3932 |
| War | Syria | 2011-24 | War | Phys | 7482* | 22.7 | 280 | 617910 |
| COVID-19 | USA | 2020-21 | Excess | Phys<br>45-84<br>years<br>old | 645427 | 330.0 | 309 | 871275 |
| COVID-19 | Ontario | 2020-22 | COVID-19 | Phys | 30617 | 14.8 | 4** | 15705 |
| COVID-19 | Finland | 2020-6/21 | COVID-19 | All | 446432 | 5.5 | 7^ | 985 |
| COVID-19 | Indonesia | 2020-7/21 | COVID-19 | All | 905261 | 271.1 | 1545 | 404650# |

HW: healthcare workers; Phys: physicians
\* the number of physicians pre-crisis (n=29927) is divided by 4 in HPH calculations
assuming the effective number during the crisis was only one quarter of the pre-crisis
numbers, as physicians massively fled the country; the correction factor nevertheless carries very large uncertainty.
\*\* 7 deaths reported within 90 days of COVID-19 infection, but 4 within 30 days; the latter is used in the HPH calculations.
^ 9 deaths in the National Infectious Diseases register, but two of them were not related to COVID-19 according to physician notifications.

### HPH estimates

Table 2 presents the absolute risk of the fatal outcome of interest in the healthcare worker group and the general population in each example, along with calculated estimates of HPH(r) and HPH(a). Absolute risks for the healthcare worker groups varied widely, from 92 Ebola deaths among 1000 healthcare workers in Sierra Leone to 0.0 COVID-19 deaths per 1000 healthcare workers in Finland. For the general population, the absolute risk spread was approximately 100-fold (0.2 to 22.4 deaths per 1000), but excluding the Syria armed conflict, the other death rates were in a more narrow range (0.2 to 2.6 per 1000).

**Table 2.**
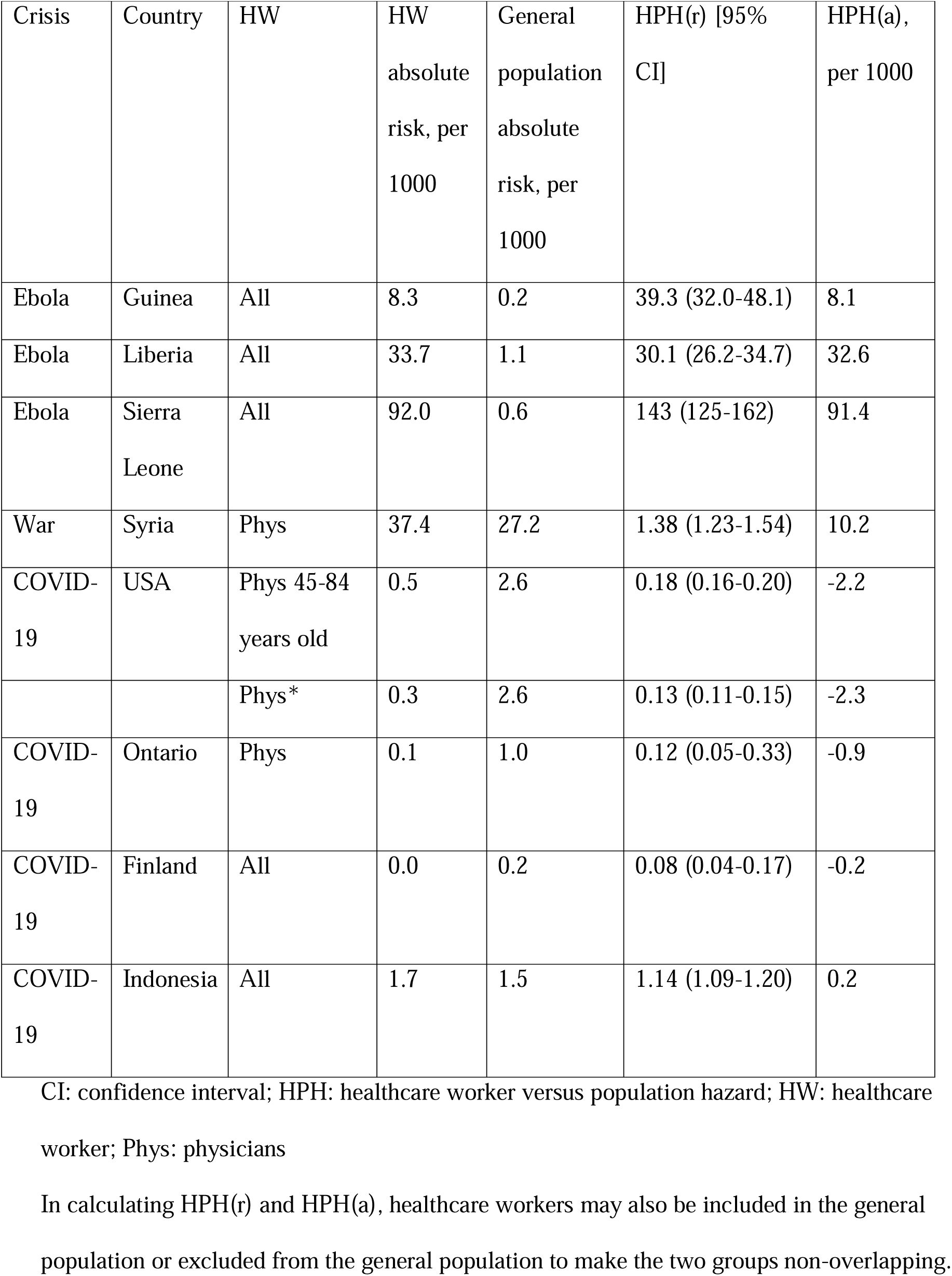
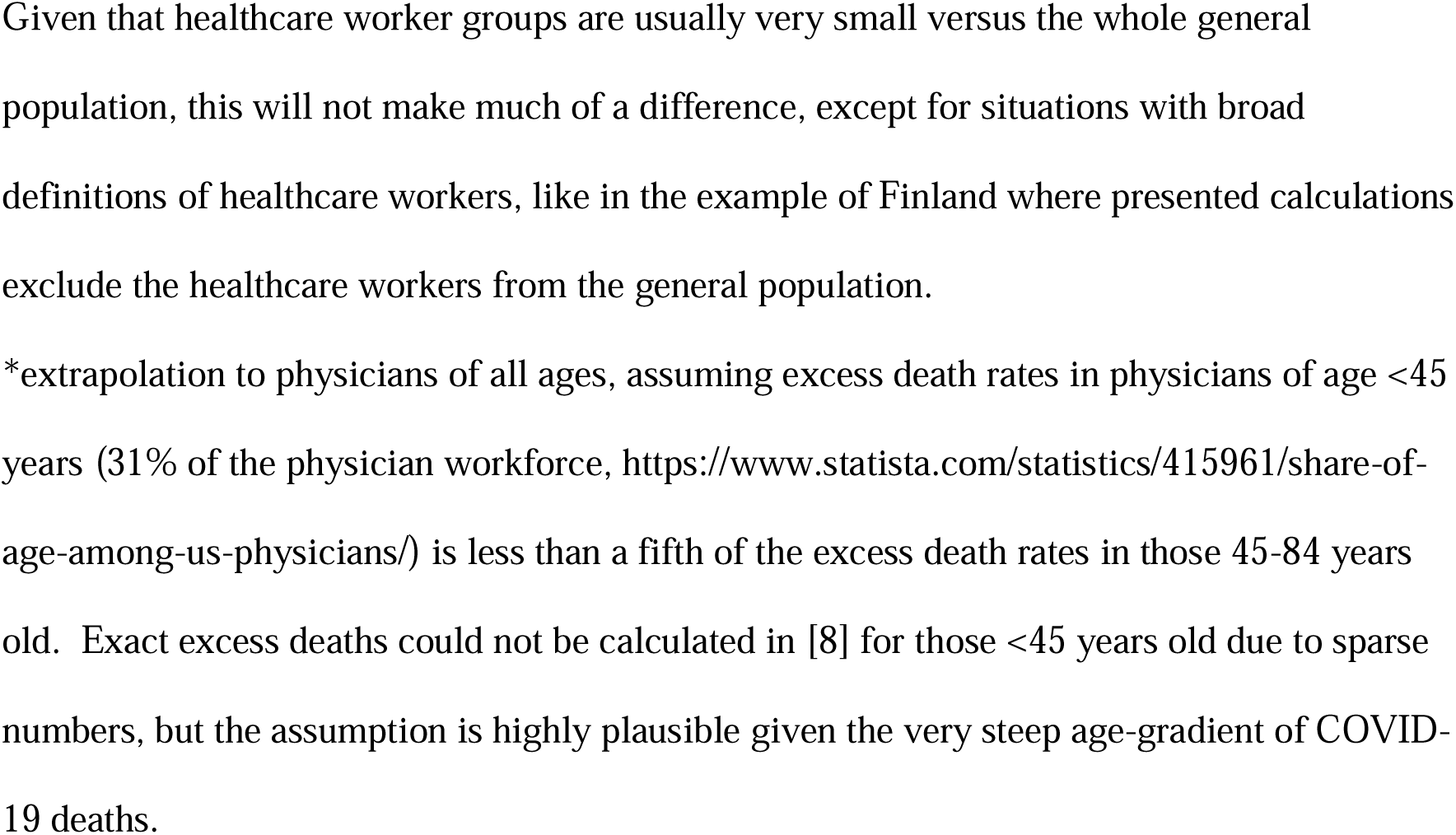
Calculation of HPH metrics for different crises.

| Crisis | Country | HW | HW<br>absolute<br>risk, per<br>1000 | General<br>population<br>absolute<br>risk, per<br>1000 | HPH(r) [95%<br>CI] | HPH(a),<br>per 1000 |
| --- | --- | --- | --- | --- | --- | --- |
| Ebola | Guinea | All | 8.3 | 0.2 | 39.3 (32.0-48.1) | 8.1 |
| Ebola | Liberia | All | 33.7 | 1.1 | 30.1 (26.2-34.7) | 32.6 |
| Ebola | Sierra<br>Leone | All | 92.0 | 0.6 | 143 (125-162) | 91.4 |
| War | Syria | Phys | 37.4 | 27.2 | 1.38 (1.23-1.54) | 10.2 |
| COVID-19 | USA | Phys 45-84<br>years old | 0.5 | 2.6 | 0.18 (0.16-0.20) | -2.2 |
|  |  | Phys* | 0.3 | 2.6 | 0.13 (0.11-0.15) | -2.3 |
| COVID-19 | Ontario | Phys | 0.1 | 1.0 | 0.12 (0.05-0.33) | -0.9 |
| COVID-19 | Finland | All | 0.0 | 0.2 | 0.08 (0.04-0.17) | -0.2 |
| COVID-19 | Indonesia | All | 1.7 | 1.5 | 1.14 (1.09-1.20) | 0.2 |

CI: confidence interval; HPH: healthcare worker versus population hazard; HW: healthcare worker; Phys: physicians
In calculating HPH(r) and HPH(a), healthcare workers may also be included in the general population or excluded from the general population to make the two groups non-overlapping.
Given that healthcare worker groups are usually very small versus the whole general population, this will not make much of a difference, except for situations with broad definitions of healthcare workers, like in the example of Finland where presented calculations exclude the healthcare workers from the general population.
\*extrapolation to physicians of all ages, assuming excess death rates in physicians of age <45 years (31% of the physician workforce, https://www.statista.com/statistics/415961/share-of-age-among-us-physicians/) is less than a fifth of the excess death rates in those 45-84 years old. Exact excess deaths could not be calculated in [8] for those <45 years old due to sparse numbers, but the assumption is highly plausible given the very steep age-gradient of COVID-19 deaths.

The relative hazard HPH(r) was extreme for Ebola (30-143 higher risk of death than the general population), and modestly high for the Syria armed conflict (1.4-fold higher risk). For COVID-19, in USA, Ontario and Finland the risk was 8-12-fold lower in healthcare workers (physicians in USA and Ontario, all healthcare workers in Finland) versus the general population. In Indonesia healthcare workers had minimally higher death risk than the general population, but the differential could be substantially greater, if healthcare worker COVID-19 deaths had also been undercounted (e.g. HPH(r)=2.28 and 3.42 and HPH(a)=1.9 and 3.6, in sensitivity analyses with 2- and 3-fold undercounting, respectively).

The absolute differential hazard HPH(a) was extremely high for Ebola and varied markedly across the 3 affected countries (8 to 91 per 1000). It was also very large in Syria (10 per 1000), and showed substantially favorable outcomes in the COVID-19 crisis for healthcare workers in the 3 developed country locations (USA, Ontario, Finland), but not in Indonesia.

Supplementary Table shows illustratively estimates of HPH(r) and HPH(a) for each year in Syria. Data should be seen with major caution given the large uncertainties about missingness and the accuracy of corrections (see Methods).

In illustrative age- and direct care-stratified analyses, active physicians in the USA who were younger than 75 years old had a large advantage over the general population they served as they had lower excess deaths regardless of whether they were involved or not in direct patient care (Table 3). In physicians who were 45-64 years old and had no direct patient care the point estimate suggested even a death deficit (fewer deaths in the pandemic versus the pre-pandemic years). Among active physicians 75-84 years old, those who had direct patient care had a modestly higher risk than the general population, while those without direct patient care had a modestly lower risk than the general population with absolute risk differences in the range of 1 in 1000.

**Table 3.** HPH for excess deaths among active physicians versus the general USA population in strata defined by age and provision of direct patient care.

| Age of physicians (years) | Direct patient care | Active physicians | Excess deaths in Active physicians | HPH(r) | HPH(a) per thousand |
| --- | --- | --- | --- | --- | --- |
| 45-64 | Yes | 426015 | 81 | 0.07 | -2.5 |
| 45-64 | No | 37648 | 13 | 0.13 | -2.3 |
| 65-74 | Yes | 133743 | 108 | 0.31 | -1.8 |
| 65-74 | No | 16229 | -8 | 0* | -3.1 |
| 75-84 | Yes | 25524 | 95 | 1.41 | 1.1 |
| 75-84 | No | 6268 | 10 | 0.60 | -1.0 |

HPH: healthcare worker versus population hazard
For details on the data, see reference [8].
\*death deficit in these physicians (fewer deaths during the pandemic years versus the pre-pandemic years)

## DISCUSSION

HPH metrics allow comparative risk assessments for active healthcare workers and general populations during crises. As shown, these metrics can vary enormously. They can reach extreme values in both relative and absolute terms during an outbreak like Western African Ebola where largely unprotected healthcare workers in understaffed facilities were exposed far more than the average citizen. They can be very high in absolute terms for healthcare workers who persevere offering their services in war-torn areas. Conversely, for COVID-19, HPH(r) in high-income countries was very low and HPH(a) was negative, suggesting that physicians or even all healthcare workers in such countries had much lower risk of COVID-19 deaths and lower excess deaths overall than the general population that they served. This privilege may not exist nevertheless in less developed countries.

Prior experience from past crises can be extrapolated to new, similar emerging crises. New data collected also in real time can help recalibrate HPH metrics, monitor their evolution and assess the impact of interventions. E.g. an Ebola outbreak may have by default a higher death rate among healthcare workers than the average person in the community. However, this grim situation can be ameliorated with higher availability of protective measures for healthcare workers, vaccination (given the current availability of apparently effective vaccine options [26]), and processes that minimize unnecessary exposures.

During wars, a main problem is the attacks on healthcare professionals that constitute an organized program of terror [7,15]. In Syria, most healthcare worker deaths were even caused by the government and its allies [15]. It is important to monitor atrocities, identify perpetrators, and intervene to avert further escalation. Accurate data collection in war conditions becomes very difficult and it may also be subverted, suppressed, or fabricated. However, some patterns of excessive risk for targeted healthcare workers may be common across very different countries and types of war conflicts. A literature of such observations has been amassed from conflicts in other areas such as Sudan, Pakistan, Burma, Iraq, Afghanistan, Nepal, Yemen, and Ukraine [27–35].

Some calculated HPH metrics were extreme, highlighting the inordinate risks that healthcare workers are occasionally exposed to. The highest absolute death risk (92 per 1000, i.e. a literal decimation) and HPH(a) (91 per 1000) was seen in the Sierra Leone Ebola outbreak. Sierra Leone had the lowest availability of healthcare workers (<0.04 per 1000 population) [14]. If data on the number of healthcare workers are accurate, the volume of Ebola cases handled per worker was also higher than elsewhere, leading to devastating outcomes for this highly vulnerable workforce. More generally, professionals working in understaffed healthcare systems may face disproportional risk, besides the burnout that is highly prevalent even in more developed countries [36]. In a vicious circle, understaffed systems have great difficulty to retain healthcare workers and witness large attrition rates [37]. Qualified physicians are sparse in these settings and newly trained physicians massively immigrate [38,39].

Conversely, for some types of crises, healthcare workers may be at lower risk than the people they serve (8,18,21). Their absolute risk may be small or even non-existent. COVID-19 was a classic example in this regard. When the pandemic started there was widespread fear of decimated healthcare workers, overrun hospitals and system collapse. This outlook contributed major psychological distress to healthcare workers [40]. Fear of COVID-19 was a major contributing theme for healthcare workers’ intention to quit [41]. The same outlook probably also facilitated the adoption of aggressive population-wide policy measures [42,43]. Nevertheless, in most developed countries, physicians and even other healthcare workers may have suffered minimal excess deaths, if any. Kiang et al. found far lower excess deaths in physicians versus the general population even after adjusting for age and gender [8]. Therefore this “privileged” low-risk status was conferred primarily by factors other than just demographics, e.g. a healthy worker effect [44], better health/lifestyle choices [45] and (by definition) higher socioeconomic status. Higher socioeconomic status strongly correlates with higher life expectancy in general [46] and low socioeconomic position was associated with 5-fold higher COVID-19 deaths [47]. Healthcare workers often had also better access to and use of effective treatments and vaccines. Finally, many, perhaps most, healthcare workers were infected in the community rather than in occupational settings; seroprevalence rates between healthcare workers and the general population usually did not markedly diverge [48].

Physicians had minimal excess deaths even in USA that had the highest excess deaths proportion among non-elderly individuals than any other country with reliable death registration [49]. Ontario (that had much lower general population excess deaths than USA), physicians also had ∼8 times lower COVID-19 deaths than the general population. Across Canada, until Jan 14, 2022, healthcare workers accounted for only 46 of 30,756 reported COVID-19 deaths [50]. While healthcare worker definitions are not sufficiently consistent in these data to allow accurate calculations, HPH(r) in Canada may have been even lower than Ontario. For countries (such as in the Nordic Region) and provinces that had even lower excess deaths of even a death deficit in the pandemic versus the pre-pandemic period [49], healthcare workers may have fared even better. In Finland, data on all healthcare workers use an expanded definition of health and social work and 8% of the total population are included in this occupational bracket; among them the risk of COVID-19 death in 2020-21 was 10 times lower than the general population [21]. In British Columbia, a province of 5 million inhabitants, during 2020-2021 not a single healthcare worker died of COVID-19 among ∼11 thousand infected [39]. Conversely, this privileged status may not exist for some low-pay, disadvantaged healthcare workers in some developed countries and for the majority of healthcare workers in less developed countries. There are large uncertainties in death ascertainment in less developed countries [51]. However, their healthcare workers probably suffered the same fate as the general population, or even worse.

Making inferences on healthcare workers from the general population experience becomes misleading if HPH is ignored. A WHO report covering COVID-19 deaths until May 2021 [52] assumed that healthcare workers had the same COVID-19 death rate as the general population in each country. It thus estimated they must have suffered 115,500 COVID-19 deaths or even 179,500 after adjusting for under-reporting – versus 6,633 officially reported deaths. However, most of these speculated deaths were imputed in developed countries; as shown here, modeling on the general population may have led to major (5-10-fold) overestimation. The same report [52] estimated 39,875 COVID-19 deaths among healthcare workers with a different method, assuming 0.4% infection fatality rate, which is probably also substantially exaggerated for an active, non-elderly population [53].

Death risk estimates and perceptions can be markedly inflated if retired healthcare workers are included in the calculations. This was a major misleading factor in early pandemic media stories and registries of physician deaths. Deaths pertaining mostly to elderly retirees were extrapolated to the active workforce. The most dramatic early data on physician deaths came from Italy, where the national registry included retirees. Few of these deaths reflected active physicians [54]. As a group, retirees largely followed the fates of high-risk general population subsets. Actually, often nursing homes were even disproportionately hit in the early pandemic [55] and high-risk people were unfortunately less protected than low-risk individuals [56]. Misclassifications of retiree deaths as active occupational risk gave rise to stunning misunderstandings, ranging from passionate editorials by esteemed opinion leaders [57] to extremely exaggerated estimates of fatality rates [58]. E.g., a systematic review of healthcare worker deaths in 2020 [58] found 37.2% fatality rate among those >=70 years old – massively inflated due to missed infections and inclusion of retirees. Infection fatality rates in active practitioners in this age range were probably 10-50 times lower [59]. In the examples analyzed in the current paper, contamination with retiree data is probably low. However, if any such misclassification ensued, then HPH is even lower than the low values obtained here.

Several limitations regarding HPH metrics should be discussed. Calculating HPH metrics is limited by data inaccuracy in outcomes and populations at-risk. For several crises, even deaths are extensively miscounted [6]. Even to obtain a simple trend, the reliability of the data needs to be verified. Healthcare professionals may have health-related data of better quality and lower missingness than the average person. This possibility should be examined on a case-by-case basis in different circumstances. The ability to have a biological diagnosis of an infection or even a clinical one may be different in the general population than in healthcare workers. This may be particularly true in developing countries, but it may also be seen in developed countries, especially in settings and populations with limited resources and suboptimal access to healthcare. In war-torn countries, population-at-risk numbers may be unknown and volatile, affected by refugee status, displacement and incarceration. In each case, one should scrutinize carefully the quality and reliability of previously available data, identify gaps and major sources of bias in data collection and reporting and try to fill some of these gaps or use more appropriate adjustments and correction factors in the future. Given the difficulty to correct fragmented data, HPH metrics should be interpreted with sufficient uncertainty surrounding them.

Finally, different healthcare workers may have different HPH metrics. Here, for illustrative purposes only age strata and direct patient care were explored, but sometimes additional stratifications may be of interest. More major differences may arise if the population served is stratified. E.g., in developed countries children and adolescents typically had a substantial death deficit during the COVID-19 pandemic versus pre-pandemic years [17,49]. It is unknown whether pediatricians suffered any excess deaths of had an equally large or even larger death deficit. Conversely, for professionals serving nursing home facilities during the pandemic, their gap versus the nursing home residents was much wider. Also for outbreak- and war-stricken countries, HPH may vary in different parts of the country, with highest values at crisis epicenters.

Allowing for these caveats, HPH may be a useful concept to measure, monitor and compare across settings and both within and between different crises. It also offers a reminder of the sacrifices demanded of healthcare workers worldwide, often under very adverse circumstances.

## Data Availability

All data produced in the present work are contained in the manuscript

## Statements and declarations

### Funding

The work of John Ioannidis is supported by an unrestricted gift from Sue and Bob O’Donnell to Stanford University.

### Data sharing

All relevant data are in the manuscript and supplement.

### Competing interests

I have worked for several years in a virology laboratory with BL3 practices handling viruses that had no meaningfully effective treatment at the time, but not specifically with Ebola. I have not practiced medicine in war-torn areas. I have not practiced clinical medicine in the USA since I moved to Stanford in 2010, but I am still actively board certified/licensed in Greece and I was consulted as an infectious disease expert on a large number of COVID-19 patients in Greece, whenever I was physically there and also remotely. I have not accepted any renumeration for any clinical work that I have provided since I finished my infectious diseases clinical fellowship training in 1996. I have witnessed many colleagues who were mentally devastated during the COVID-19 pandemic, terrorized by misinformation that stressed their supposedly high risks of death and other adverse outcomes from SARS-CoV-2. This article is dedicated to all physicians, other healthcare workers and researchers who expose themselves to high risks to serve humanity and to those who burned out, in part because they were misinformed about the risks they face.

### Author contributions

JPAI conceived the concept, collected data, analyzed data, wrote the manuscript and approved the final version.

### Ethics approval

Not relevant (no new data collected, methodological work).

### Supplementary text

For healthcare workers other than physicians, the proportion who fled the country is also uncertain, but is likely to have been also substantial. The illustrative calculations assumed that total healthcare workers may have been close to 2.5-fold the number of physicians before the crisis, but dropped to 60 thousand during 2011 and then there was a 30% reduction for 2012, another 20% reduction for 2013, 10% reduction for 2014, 10% reduction for 2015 and stabilization afterwards at 18 thousand (30% of the 2011 level).

**Supplementary Table.** Approximation of HPH for each calendar year between 2011 and 2021 in the Syrian armed conflict.

| Year | HW | Pop<br>(thousands) | HW<br>deaths<br>from<br>war | Pop deaths<br>from war | HPH(r) | HPH(a)<br>per<br>thousand |
| --- | --- | --- | --- | --- | --- | --- |
| 2011 | 60000 | 22731 | 52 | 7841 | 2.5 | 0.5 |
| 2012 | 42000 | 22606 | 190 | 52916 | 1.9 | 2.2 |
| 2013 | 30000 | 21496 | 180 | 97009 | 1.3 | 1.5 |
| 2014 | 24000 | 20072 | 176 | 110920 | 1.3 | 1.8 |
| 2015 | 18000 | 19205 | 108 | 85527 | 1.3 | 1.5 |
| 2016 | 18000 | 18964 | 91 | 64126 | 1.5 | 1.7 |
| 2017 | 18000 | 18983 | 51 | 38202 | 1.4 | 0.8 |
| 2018 | 18000 | 19333 | 45 | 20133 | 2.4 | 1.5 |
| 2019 | 18000 | 20098 | 26 | 11244 | 2.6 | 0.9 |
| 2020 | 18000 | 20773 | 9 | 6762 | 1.5 | 0.2 |
| 2021 | 18000 | 21324 | 11 | 3884 | 3.4 | 0.4 |

HPH: healthcare worker versus population hazard; HW: healthcare worker; Pop: population.

Number of deaths are small after 2021 and not shown here. All estimates carry very large uncertainty. Recorded war-related deaths may be undercounted both for HW and the general population and the evolution of HW numbers over time is very uncertain (see Methods).

## Notes

### Competing Interest Statement

The authors have declared no competing interest.

### Funding Statement

The study did not receive any funding

### Summary of Updates

clarifications in methods; elaboration of discussion on war; addition of some limitations; editing

## REFERENCES

[1] Sepkowitz KA, Eisenberg L. Occupational deaths among healthcare workers. Emerg Infect Dis. 2005 Jul;11(7):1003–8.

[2] Sepkowitz KA. Occupationally acquired infections in health care workers. Part II. Ann Intern Med. 1996 Dec 1;125(11):917–28.

[3] Sepkowitz KA. Occupationally acquired infections in health care workers. Part I. Ann Intern Med. 1996 Nov 15;125(10):826–34.

[4] Haar RJ, Risko CB, Singh S, Rayes D, Albaik A, Alnajar M, Kewara M, Clouse E, Baker E, Rubenstein LS. Determining the scope of attacks on health in four governorates of Syria in 2016: Results of a field surveillance program. PLoS Med. 2018 Apr 24;15(4):e1002559.

[5] GBD 2019 Human Resources for Health Collaborators. Measuring the availability of human resources for health and its relationship to universal health coverage for 204 countries and territories from 1990 to 2019: a systematic analysis for the Global Burden of Disease Study 2019. Lancet. 2022 Jun 4;399(10341):2129–2154.

[6] Ioannidis JPA. Over- and under-estimation of COVID-19 deaths. Eur J Epidemiol. 2021 Jun;36(6):581–588. doi: 10.1007/s10654-021-00787-9.

[7] Fouad FM, Sparrow A, Tarakji A, Alameddine M, El-Jardali F, Coutts AP, El Arnaout N, Karroum LB, Jawad M, Roborgh S, Abbara A, Alhalabi F, AlMasri I, Jabbour S. Health workers and the weaponisation of health care in Syria: a preliminary inquiry for The Lancet-American University of Beirut Commission on Syria. Lancet. 2017 Dec 2;390(10111):2516–2526.

[8] Kiang MV, Carlasare LE, Thadaney Israni S, Norcini JJ, Zaman JAB, Bibbins-Domingo K. Excess Mortality Among US Physicians During the COVID-19 Pandemic. JAMA Intern Med. 2023 Apr 1;183(4):374–376.

[9] World Health Organization, Ebola situation report, July 1, 2015.

[10] Grinnell M, Dixon MG, Patton M, Fitter D, Bilivogui P, Johnson C, Dotson E, Diallo B, Rodier G, Raghunathan P. Ebola Virus Disease in Health Care Workers--Guinea, 2014. MMWR Morb Mortal Wkly Rep. 2015 Oct 2;64(38):1083–7.

[11] Kilmarx PH, Clarke KR, Dietz PM, Hamel MJ, Husain F, McFadden JD, Park BJ, Sugerman DE, Bresee JS, Mermin J, McAuley J, Jambai A; Centers for Disease Control and Prevention (CDC). Ebola virus disease in health care workers—Sierra Leone, 2014. MMWR Morb Mortal Wkly Rep. 2014 Dec 12;63(49):1168–71.

[12] Liberia Malaria Impact Evaluation Group. Evaluation of the Impact of Malaria Control Interventions on All-Cause Mortality in Children Under Five Years of Age in Liberia, 2005– 2013, USAID 2018.

[13] Wordometer, 2013 populations. In: https://www.worldometers.info/world-population/world-population-countries.php, last accessed May 23, 2024.

[14] Syrian Observatory for Human Rights. https://www.syriahr.com/en/328044/, last accessed May 23, 2024.

[15] Physicians for Human Rights, https://phr.org/our-work/resources/medical-personnel-are-targeted-in-syria/, last accessed May 23, 2024.

[16] Population of Syria. "World Population Prospects - Population Division - United Nations". In: population.un.org. Last accessed May 24, 2024.

[17] Levitt M, Zonta F, Ioannidis JPA. Comparison of pandemic excess mortality in 2020-2021 across different empirical calculations. Environ Res. 2022 Oct;213:113754.

[18] Habbous S, Saunders N, Chan KK, Hota S, Wang J, Messenger D, Hellsten E. SARS-CoV-2 infection among physicians over time in Ontario, Canada: a population-based retrospective cohort study. Croat Med J. 2024 Feb 29;65(1):30-42.

[19] Ontario Demographic Quarterly Highlights. In: https://www.ontario.ca/page/ontario-demographic-quarterly-highlights-third-quarter, last accessed May 23, 2024.

[20] . Health Infobase. In: https://health-infobase.canada.ca/covid-19/archive/2023-01-02/#a3. Last accessed May 24, 2024.

[21] Kääriäinen S, Harjunmaa U, Hannila-Handelberg T, Ollgren J, Lyytikäinen O. Risk of COVID-19 in different groups of healthcare professionals between February 2020 and June 2021 in Finland: a register-based cohort study. Infect Prev Pract. 2023 Jul 6;5(3):100297.

[22] Worldometer. In: https://www.worldometers.info/world-population/finland-population/, last accessed May 24, 2024.

[23] Coronavirus in Finland. In: https://www.worldometers.info/coronavirus/country/finland/, last accessed May 24, 2024.

[24] Ekawati LL, Arif A, Hidayana I, Nurhasim A, Munziri MZ, Lestari KD, Tan A, Ferdiansyah F, Nashiruddin F, Adnani QES, Malik H, Maharani T, Riza A, Pasaribu M, Abidin K, Andrianto AA, Nursalam N, Suhardiningsih AVS, Jubaedah A, Widodo NS, Surendra H, Sudoyo H, Smith AD, Kreager P, Baird JK, Elyazar IRF. Mortality among healthcare workers in Indonesia during 18 months of COVID-19. PLOS Glob Public Health. 2022 Dec 9;2(12):e0000893.

[25] IHME, Indonesia COVID-19 data and projections. In: https://covid19.healthdata.org/indonesia?view=cumulative-deaths&tab=trend, last accessed May 23, 2024.

[26] PREVAC Study Team; Kieh M, Richert L, Beavogui AH, Grund B, Leigh B, D’Ortenzio E, Doumbia S, Lhomme E, Sow S, Vatrinet R, Roy C, Kennedy SB, Faye S, Lees S, Millimouno NP, Camara AM, Samai M, Deen GF, Doumbia M, Espérou H, Pierson J, Watson-Jones D, Diallo A, Wentworth D, McLean C, Simon J, Wiedemann A, Dighero-Kemp B, Hensley L, Lane HC, Levy Y, Piot P, Greenwood B, Chêne G, Neaton J, Yazdanpanah Y. Randomized Trial of Vaccines for Zaire Ebola Virus Disease. N Engl J Med. 2022 Dec 29;387(26):2411–2424.

[27] Cone J, Duroch F: Don’t Shoot the Ambulance: Medicine in the Crossfire. 2013, US: World Policy Institute

[28] Burnham G, Lafta R, Doocy S: Doctors leaving 12 tertiary hospitals in Iraq, 2004–2007. Soc Sci Med. 2009, 69 (2): 172–177.

[29] Morikawa M: Effect of security threats on primary care access in Logar Province, Afghanistan. Med Confl Surviv. 2008, 24 (1): 59–64.

[30] Devkota B, van Teijlingen ER: “Understanding effects of armed conflict on health outcomes: the case of Nepal,”. Confl. Health. 2010, 4 (no. 1): p. 20-10.1186/1752–1505-4-20.

[31] Varley E: Targeted doctors, missing patients: obstetric health services and sectarian conflict in northern Pakistan. Soc Sci Amp Med 1982. 2009, 70 (1): 61–70.

[32] Haar RJ, Footer KH, Singh S, Sherman SG, Branchini C, Sclar J, Clouse E, Rubenstein LS. Measurement of attacks and interferences with health care in conflict: validation of an incident reporting tool for attacks on and interferences with health care in eastern Burma. Confl Health. 2014 Nov 3;8(1):23.

[33] Badri R, Dawood I. The implications of the Sudan war on healthcare workers and facilities: a health system tragedy. Confl Health. 2024 Mar 17;18(1):22.

[34] Geiger HJ, Cook-Deegan RM. The role of physicians in conflicts and humanitarian crises. Case studies from the field missions of Physicians for Human Rights, 1988 to 1993. JAMA. 1993 Aug 4;270(5):616–20.

[35] Rubin R. Physicians in Ukraine: Caring for Patients in the Middle of a War. JAMA. 2022 Apr 12;327(14):1318–1320.

[36] Rotenstein LS, Torre M, Ramos MA, Rosales RC, Guille C, Sen S, Mata DA. Prevalence of Burnout Among Physicians: A Systematic Review. JAMA. 2018 Sep 18;320(11):1131–1150.

[37] Castro Lopes S, Guerra-Arias M, Buchan J, Pozo-Martin F, Nove A. A rapid review of the rate of attrition from the health workforce. Hum Resour Health. 2017 Mar 1;15(1):21.

[38] Cometto G, Tulenko K, Muula AS, Krech R. Health workforce brain drain: from denouncing the challenge to solving the problem. PLoS Med. 2013;10, e1001514.

[39] Burch VC, McKinley D, van Wyk J, et al. Career intentions of medical students trained in six sub-Saharan African countries. Educ Health (Abingdon). 2011;24:614.

[40] Arias-Ulloa CA, Gómez-Salgado J, Escobar-Segovia K, García-Iglesias JJ, Fagundo-Rivera J, Ruiz-Frutos C. Psychological distress in healthcare workers during COVID-19 pandemic: A systematic review. J Safety Res. 2023 Dec;87:297–312.

[41] Poon YR, Lin YP, Griffiths P, Yong KK, Seah B, Liaw SY. A global overview of healthcare workers’ turnover intention amid COVID-19 pandemic: a systematic review with future directions. Hum Resour Health. 2022 Sep 24;20(1):70

[42] Schippers MC, Ioannidis JPA, Joffe AR. Aggressive measures, rising inequalities, and mass formation during the COVID-19 crisis: An overview and proposed way forward. Front Public Health. 2022 Aug 25;10:950965.

[43] Joffe AR. COVID-19: Rethinking the Lockdown Groupthink. Front Public Health. 2021 Feb 26;9:625778.

[44] Li CY, Sung FC. A review of the healthy worker effect in occupational epidemiology. Occup Med (Lond). 1999 May;49(4):225–9.

[45] Frank E, Brogan DJ, Mokdad AH, et al. Health-related behaviors of women physicians vs other women in the United States. Arch Intern Med. 1998;158:342–348.

[46] Dwyer-Lindgren L, Bertozzi-Villa A, Stubbs RW, Morozoff C, Mackenbach JP, van Lenthe FJ, Mokdad AH, Murray CJL. Inequalities in Life Expectancy Among US Counties, 1980 to 2014: Temporal Trends and Key Drivers. JAMA Intern Med. 2017 Jul 1;177(7):1003–1011.

[47] Pathak EB, Menard JM, Garcia RB, Salemi JL. Joint Effects of Socioeconomic Position, Race/Ethnicity, and Gender on COVID-19 Mortality among Working-Age Adults in the United States. Int J Environ Res Public Health. 2022 Apr 30;19(9):5479.

[48] Bobrovitz N, Arora RK, Cao C, Boucher E, Liu M, Donnici C, Yanes-Lane M, Whelan M, Perlman-Arrow S, Chen J, Rahim H, Ilincic N, Segal M, Duarte N, Van Wyk J, Yan T, Atmaja A, Rocco S, Joseph A, Penny L, Clifton DA, Williamson T, Yansouni CP, Evans TG, Chevrier J, Papenburg J, Cheng MP. Global seroprevalence of SARS-CoV-2 antibodies: A systematic review and meta-analysis. PLoS One. 2021 Jun 23;16(6):e0252617.

[49] Ioannidis JPA, Zonta F, Levitt M. Variability in excess deaths across countries with different vulnerability during 2020-2023. Proc Natl Acad Sci U S A 2023 Dec 5;120(49):e2309557120.

[50] COVID-19 cases and deaths in healthcare workers in Canada. In: https://www.cihi.ca/en/covid-19-cases-and-deaths-in-health-care-workers-in-canada-infographic, last accessed May 24, 2024.

[51] Ioannidis JPA, Zonta F, Levitt M. Flaws and uncertainties in pandemic global excess death calculations. Eur J Clin Invest. 2023 Aug;53(8):e14008.

[52] World Health Organization. (2021). The impact of COVID-19 on health and care workers: a closer look at deaths. World Health Organization. https://iris.who.int/handle/10665/345300.s

[53] Pezzullo AM, Axfors C, Contopoulos-Ioannidis DG, Apostolatos A, Ioannidis JPA. Age-stratified infection fatality rate of COVID-19 in the non-elderly population. Environ Res. 2023 Jan 1;216(Pt 3):114655.

[54] Modenese A, Loney T, Gobba F. COVID-19-Related Mortality amongst Physicians in Italy: Trend Pre- and Post-SARS-CoV-2 Vaccination Campaign. Healthcare (Basel). 2022 Jun 24;10(7):1187.

[55] International Long Term Care Policy Network. Mortality associated with COVID-19 in care homes: international evidence. Available: https://ltccovid.org/2020/04/12/mortality-associated-with-covid-19-outbreaks-in-care-homes-early-international-evidence/#:~:text=Based%20on%20the%20data%20gathered,(based%20on%2021%20countries [Accessed May 23, 2024]

[56] Ioannidis JPA. Precision shielding for COVID-19: metrics of assessment and feasibility of deployment. BMJ Glob Health. 2021 Jan;6(1):e004614.

[57] Verghese A, Topol E. Courage in a climate of fear. Sci Transl Med. 2020 Nov 4;12(568):eabf2461.

[58] Bandyopadhyay S, Baticulon RE, Kadhum M, Alser M, Ojuka DK, Badereddin Y, Kamath A, Parepalli SA, Brown G, Iharchane S, Gandino S, Markovic-Obiago Z, Scott S, Manirambona E, Machhada A, Aggarwal A, Benazaize L, Ibrahim M, Kim D, Tol I, Taylor EH, Knighton A, Bbaale D, Jasim D, Alghoul H, Reddy H, Abuelgasim H, Saini K, Sigler A, Abuelgasim L, Moran-Romero M, Kumarendran M, Jamie NA, Ali O, Sudarshan R, Dean R, Kissyova R, Kelzang S, Roche S, Ahsan T, Mohamed Y, Dube AM, Gwini GP, Gwokyala R, Brown R, Papon MRKK, Li Z, Ruzats SS, Charuvila S, Peter N, Khalidy K, Moyo N, Alser O, Solano A, Robles-Perez E, Tariq A, Gaddah M, Kolovos S, Muchemwa FC, Saleh A, Gosman A, Pinedo-Villanueva R, Jani A, Khundkar R. Infection and mortality of healthcare workers worldwide from COVID-19: a systematic review. BMJ Glob Health. 2020 Dec;5(12):e003097.

[59] Axfors C, Ioannidis JPA. Infection fatality rate of COVID-19 in community-dwelling elderly populations. Eur J Epidemiol. 2022 Mar;37(3):235–249.

